# MosCoverY: a new method to estimate mosaic Loss of Y chromosome from NGS sequencing coverage data

**DOI:** 10.1101/2024.11.11.24317095

**Authors:** V. Timonina, A. Marchal, L. Abel, A. Cobat, J. Fellay

## Abstract

Mosaic loss of the Y chromosome (mLOY) is the most common somatic event in men, strongly associated with aging and various health conditions. Current methods for detecting mLOY primarily rely on DNA genotyping arrays. Here, we present MosCoverY, a novel method for estimating mLOY from NGS sequencing data that can be applied to both exome and genome sequencing. MosCoverY addresses the challenges posed by the structure of the Y chromosome by focusing on single-copy genes and normalizing their coverage against autosomal exons matched by length and GC content. We validated MosCoverY using data from 212,062 male participants in the UK Biobank, comparing its results to those obtained using genotyping- or whole genome sequencing-based methods. MosCoverY identified mLOY in 5.6% of men, demonstrating performance that was comparable to the other methods. MosCoverY also replicated known associations between mLOY, age, smoking, all-cause mortality, and germline genetic loci, showing the strongest associations in many cases. MosCoverY offers a valuable tool for detecting mLOY from exome data in population-scale studies.

## INTRODUCTION

Somatic mutations are non-inheritable changes in the DNA of somatic cells leading to genetic mosaicism. These mutations are particularly well-described in blood, where they can cause clonal hematopoiesis (1–7). The most commonly occurring somatic event in leukocytes of male individuals is the mosaic loss of chromosome Y (mLOY) (8–12). The prevalence of mLOY increases with age and is associated with various health-related conditions, including all-cause mortality (13, 14), hematological cancer (15–18), solid tumors (14, 19), cardiovascular diseases (8, 9, 20, 21), and Alzheimer’s disease (15, 22).

mLOY can be detected using targeted quantitative PCR assay, genotyping arrays or next-generation sequencing. The most commonly used methods utilize genotyping array data to detect the deviation in the intensity probe values on the Y chromosome (chrY). One such approach, mLRRY, calculates the median log-R ratio (mLRR, normalized intensity) in the male-specific part of the chrY (MSY), excluding pseudoautosomal regions (PARs), and considers values below a chosen threshold as evidence of mLOY (11, 19). Another method, PAR-LOY, uses long-range phasing information in the PARs to evaluate the allelic intensities imbalance in heterozygous sites between sex chromosomes (23).

It is also possible to estimate mLOY from whole genome sequencing (WGS) data using tools developed for copy-number variant calling in tumor cells, even if a normal sample is not available. One example is Control-FREEC, which calculates the ploidy of genomic regions from coverage data, taking into account GC content and genome mappability information (24, 25). Although the mLOY estimation from Control-FREEC can be of high quality, WGS data are not always available and computationally expensive to work with. Control-FREEC can be run on exome sequencing data, but this requires a matched normal sample, similar to other copy-number callers like FACETs (8, 26).

Currently, no method is available to call mLOY from exome sequencing data that can be applied to studies with only one sample per individual. Calling mLOY from exome sequencing data presents several challenges due to limited coverage of the Y chromosome (exome capture kits target mainly coding regions), biases in GC content that can affect coverage estimation, and the presence on chrY of repetitive sequences and regions highly similar to the X chromosome (27, 28).

Here, we propose an approach to estimating mLOY from exome sequencing data that overcomes these challenges. We also show that our method can be directly used on genome sequencing data. We demonstrate its validity by comparing it with two methods based on genotyping array data - mLRRY and PAR-LOY, as well as with one WGS-based method - Control-FREEC.

## METHODS

### Study participants

We selected participants from the UK Biobank (UKB) who satisfied the following criteria: male genetic and self-reported sex, absence of sex aneuploidy, and availability of exome sequencing data. The UKB is a large, prospective cohort study consisting of approximately 500,000 participants from the United Kingdom, with extensive phenotypic, genetic, and imaging data collected (29, 30). We limited the choice of participants to those included in the Loh et al. study, which identified all mosaic chromosomal alterations, including loss of the Y chromosome, and returned the data to the UKB (Return #2062) (31). After applying all the filters, the total sample size was 212,062 individuals. For survival analyses and genome-wide association studies (GWAS), we included only individuals of European ancestry, reducing the sample size to 178,073.

### Selecting chromosome Y exons from the exome capture kit

Due to the structure of the MSY, we restricted the choice of genes that were used to calculate chrY coverage. We excluded genes in the ampliconic part of chrY as their copy number can vary a lot between individuals, and genes in X-transposed regions due to their high similarity to the X chromosome sequences, which could affect coverage estimation. We included only single-copy genes in the X-degenerate regions of MSY (as described in (27)), resulting in 13 genes (SRY, RPS4Y1, ZFY, AMELY, TBL1Y, USP9Y, DDX3Y, UTY, TMSB4Y, NLGN4Y, KDM5D, EIF1AY, RPS4Y2) and 184 exons. We further filtered out exons that were outliers by coverage in individuals without evidence of mLOY as identified by the PAR-LOY method, reducing the number of exons to 173 (out of 45 genes with 578 exons captured by the exome kit used by UKB; selected exons are listed in Supplementary table 1).

### Selecting autosomal exons for normalization

For each selected exon on chrY, we found 100 autosomal exons matched by GC content and length (Fig. S1). These autosomal exons were selected from the regions of the genome that are rarely gained or lost in blood (less than 10 gains and losses in the UKB database) (32; UKB Return #2062) and matched by propensity score using the *MatchIt* R library. All matched exons are listed in Supplementary table 2.

### Computing normalized chromosome Y coverage

We calculated the median coverage of each of the selected exons (on chrY and autosomes) with the *mosdepth* tool from exome alignment files in .cram format (33). We normalized the median coverage of each selected chrY exon to the median coverage of its 100 matched autosomal exons, and the median of the results was an individual-level estimate of normalized chrY coverage (Figure 1A). Normalization to matched autosomal exons significantly reduced noise and improved correlation with other methods compared to normalization to the median coverage of all exons on chromosome 1 (Fig. S2). The distribution of median chrY coverage in the population was then rescaled to 0.5, the expected value given chrY’s haploid nature.

**Figure 1.**
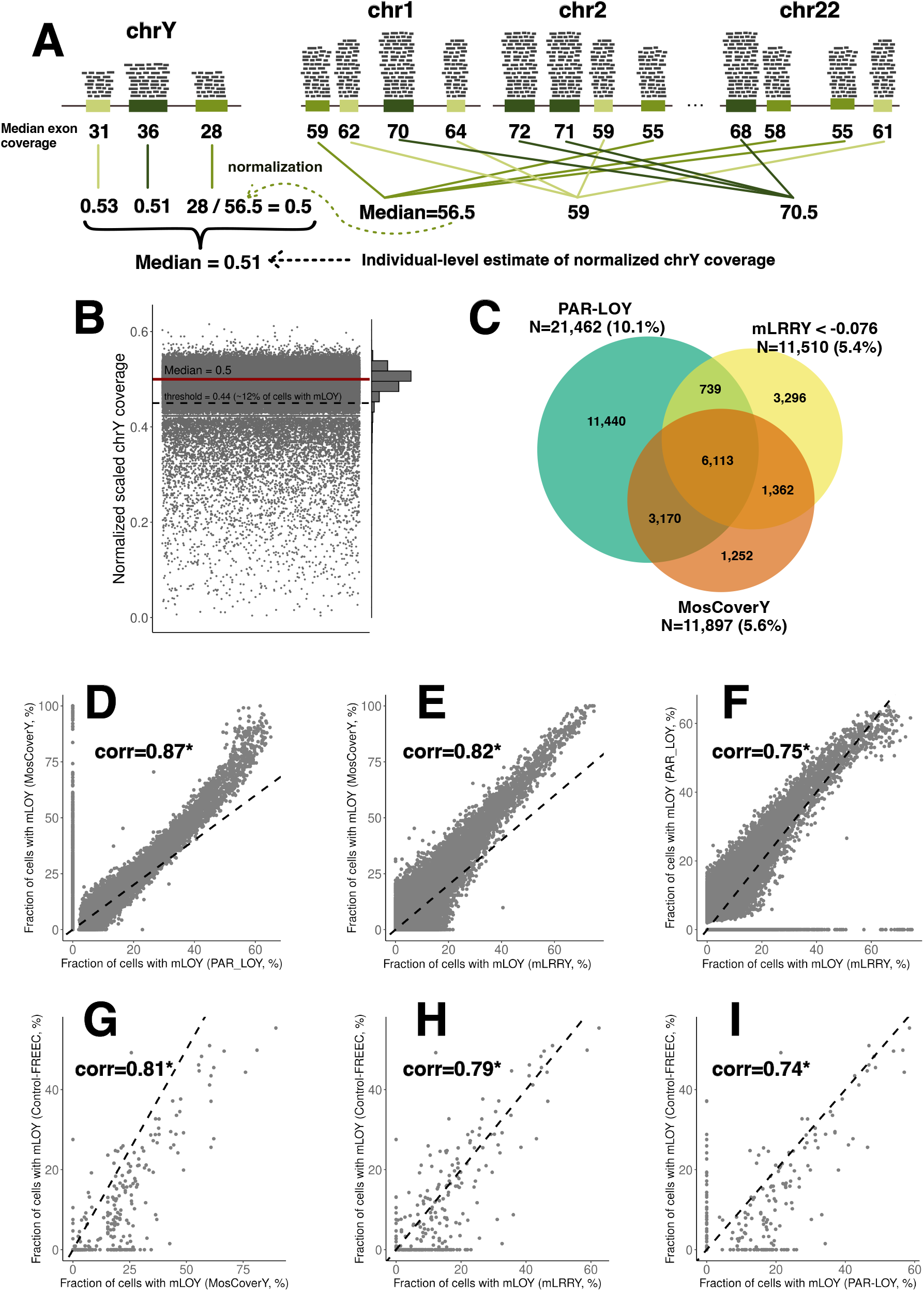
Description of the MosCoverY method and comparison with mLOY results obtained with mLRRY and PAR-LOY. A - One of the main principles of MosCoverY is a normalization of median coverage of selected chrY exons to a set of autosomal exons matched on GC content and length. Matched groups are represented by different shades of green color. The individual-level estimation of chrY coverage corresponds to the median across all normalized chrY exon coverage values. B - The distribution of normalized scaled chrY coverage in the UKB. Each dot represents an individual. The median of the distribution was scaled to 0.5 (red line), and the threshold to define mLOY was derived as Q1-1.5*IQR and is equal to 0.44 (dashed black line). All individuals with normalized scaled chrY coverage below the threshold are considered as mLOY cases. C - Venn diagram comparing MosCoverY results with two other mLOY calling methods - PAR-LOY and mLRRY - with an indication of the total number of individuals with mLOY and their proportion according to each of the methods. D, E, F - Correlation of the fraction of cells with mLOY derived from each of the three methods - MosCoverY vs. PAR-LOY, MosCoverY vs. mLRRY, and mLRRY vs. PAR-LOY. The Black dashed line represents a diagonal. (corr - Pearson’s correlation coefficient, * - p-value ≤ 0.001). G,H,I - Correlation of the fraction of cells with mLOY derived from MosCoverY, mLRRY and PAR-LOY with cell fraction estimated from WGS with Control-FREEC for a subset of 360 randomly chosen individuals.The Black dashed line represents a diagonal. (corr - Pearson’s correlation coefficient, * - p-value ≤ 0.001).

### Defining binary mLOY threshold

We defined a binary trait of mLOY by setting the threshold at Q1-1.5*IQR of the scaled chrY coverage distribution. In the UKB, this threshold was 0.44, corresponding to approximately 12% of cells with mLOY (using formula 1).

### Converting scaled normalized chrY coverage to cell fraction

We converted the scaled normalized coverage of chrY to a fraction of cells with mLOY using the following formula (Fig. S3):

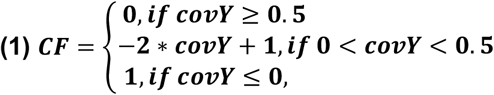

where CF - fraction of cells with mLOY; covY - normalized scaled coverage of the Y chromosome.

### Other mLOY estimation methods

To evaluate the performance of our method, we compared it to state-of-the-art methods used to estimate mLOY from genotyping or whole genome sequencing data.

The first method uses the median log-R ratio over genotyping probes on MSY (mLRRY). We calculated mLRRY over 691 SNP probes in MSY and converted mLRRY to the fraction of cells with mLOY using the formula (25):

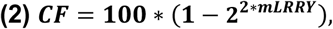

where CF - fraction of cells with mLOY; mLRRY - median log-R ratio over genotyping probes on MSY.

We used the threshold of mLRRY < -0.076 to define the binary mLOY trait, corresponding to 10% of cells with mLOY as previously published (13)).

The second method, PAR-LOY, uses genotyping array probe intensity to define allelic imbalance in heterozygous sites of PARs of chrY (Thompson et al. 2019). The PAR-LOY estimate is available as part of the UKB return #2062 and provides both the binary mLOY estimates and the fraction of cells with mLOY (which is less than 10% for the majority of people).

For a random subset of 360 individuals, we estimated relative chrY coverage with Control-FREEC software (24, 34). Control-FREEC segments the genome and identifies copy-number alterations using GC-content normalization and genome mappability information. We ran it on WGS alignment files and calculated the median ratio (normalized coverage) across all windows, excluding windows with a median ratio of more than one as they likely represent repetitive regions of chrY and would add noise to the mLOY estimation. To convert it to the cell fraction we used formula 1, as for MosCoverY.

### Epidemiological validation: Association of mLOY with age and smoking, as well as with all-cause mortality

To validate our method, we assessed the associations of mLOY with age and smoking status. We fitted a logistic regression model for the binary estimates of mLOY and a linear regression model for the fraction of cells with mLOY. We built separate models for each mLOY estimation method, fitting age at the first visit to the assessment center and ever-smoking status in the same model. We used the *glm* function with binomial family and *lm* function from R statistical software (v.4.1.1) to fit logistic and linear models, respectively.

We also performed a survival analysis of all-cause mortality, modeling time to death using multivariable-adjusted Cox proportional hazard regression with age at the first visit to the assessment center, smoking status (current, previous, or never), and genetic principal components as covariates. Time interaction terms for age and smoking status were included to satisfy the proportional hazards assumption of the Cox model. The target variable was the binary mLOY trait estimated by the various methods. We used the *coxph* function from the survival package of R statistical software (v.4.1.1).

### Genetic validation: Genome-Wide Association Study

For the Genome-Wide Association Study (GWAS) we focused on men of European ancestry. We filtered out variants with MAF less than 1%, minor allele count less than 20, missing call frequencies greater than 0.1, and strong Hardy-Weinberg equilibrium deviation (p<1e-15). Genetic principal components (PCs) for the set of European-ancestry men were calculated with plink2 on an LD-pruned set of 149,605 SNPs using the *snpgdsLDpruning* function from the *SNPRelate* R package.

We performed genetic association testing with *regenie* (Mbatchou et al., 2021) in two steps: whole-genome ridge regression to reduce the dimension of genetic data (using N=611,713 variants), followed by association testing with the larger set of variants (N=9,607,831 imputed variants). Covariates included age, ever-smoking status, genotyping batch, WES release, and 10 genetic PCs.

We identified statistically independent signals with GCTA-COJO (Genome-wide Complex Trait Analysis multi-SNP-based conditional & joint association analysis using GWAS summary data) (35, 36). We used a p-value threshold of 5e-8 to identify genome-wide significant SNPs, considering SNPs more than 1Mb apart to be in linkage equilibrium and setting the collinearity threshold between SNPs to less than 0.9.

## RESULTS

### Estimation of mLOY from Exome sequencing data

We developed a novel approach that we called MosCoverY (mosaic Loss of Y chromosome from NGS sequencing coverage data) to estimate mLOY primarily from exome sequencing data. MosCoverY uses alignment files (in .bam or .cram format) and performs several steps to estimate the normalized median coverage of chrY. Key features of MosCoverY include focusing on single-copy genes on the Y chromosome and normalizing the coverage of exons in these genes to autosomal exons matched by GC content and length. This approach provides an individual-level estimate of normalized median chrY coverage, which is then scaled to match the expected population median of 0.5 (see Methods for details). Using this normalized median coverage, we statistically derive a binary mLOY estimate as Q1-1.5*IQR, so that individuals with normalized scaled coverage below this value are classified as having mLOY (Fig. 1A).

The distribution of normalized scaled chrY coverage is shown in Figure 1B. The individuals with normalized scaled chrY coverage higher than 0.6 and lower than 0 were excluded. Using the binary threshold of 0.44 (derived as Q1-1.5*IQR), MosCoverY identified 11,897 (5.6%) men with mLOY. We converted the scaled normalized coverage of chrY to a fraction of cells with mLOY using Formula 1 (Methods). The mean fraction of cells with mLOY estimated by MosCoverY in all UKB participants was 2.8% (including individuals without mLOY and hence with 0% of cells with mLOY).

In comparison, binary estimates based on mLRRY and PAR-LOY identified 11,510 (5.4%) and 21,462 (10.1%) men with mLOY, respectively. Across all three methods, mLOY was identified in 6,113 men (22.3% of all mLOY cases, Figure 1C). Unique to MosCoverY, 1,252 men (4.6% of the union of all mLOY cases) were identified, compared to 3,296 (12%) and 11,440 (41.8%) identified solely by mLRRY and PAR-LOY, respectively. The normalized scaled coverage of chrY was on average lower for mLOY cases identified by MosCoverY only compared to those identified by either mLRRY or PAR-LOY or both, but it was higher compared to cases identified by MosCoverY together with either mLRRY or PAR-LOY or both (Fig. S4).

The mean fraction of cells with mLOY estimated by mLRRY and PAR-LOY was 2.4% and 1.3%, respectively, both lower than the 2.8% estimated by MosCoverY. The fraction of cells with mLOY estimated by MosCoverY showed a strong correlation with both mLRRY and PAR-LOY estimates (Pearson’s correlation coefficient > 0.8; Figures 1C and 1D), which was higher than the correlation between mLRRY and PAR-LOY estimates (Pearson’s correlation coefficient = 0.75; Figure 1F). We also estimated the fraction of cells with mLOY from WGS data with the Control-FREEC tool (24, 34) for a subset of 360 randomly chosen individuals. The MosCoverY-derived fraction of cells with mLOY showed better correlation with Control-FREEC-derived cell fraction as compared to mLRRY and PAR-LOY (Figure 1G-I).

Although our primary goal was to develop a method for identifying mLOY from exome sequencing data, MosCoverY can also be applied directly to WGS data. We demonstrated this by applying MosCoverY to a subset of 4,200 randomly selected individuals with genome sequencing data. The normalized coverage of chrY estimated from exome and genome sequencing showed a strong correlation (Pearson’s correlation = 0.9). Furthermore, 74% of individuals identified as mLOY cases were consistent between exome and genome sequencing (Fig. S5). Additionally, exome and genome sequencing data can be combined for mLOY identification. However, in this case they should be scaled separately before combining to define the mLOY threshold, rather than being combined prior to scaling (compare Fig. S6A and Fig. S6B). mLOY estimated from WGS data with MosCoverY also correlated well with mLOY estimated from WGS data with Control-FREEC in a subset of 360 individuals (Pearson’s correlation = 0.72 for normalized scales coverage and 0.82 for the fraction of cells with mLOY, Fig. S7).

### Epidemiological validation of MosCoverY

In the absence of a gold-standard method to assess the presence of mLOY, we aimed at validating the performance of our approach by examining and comparing the strength of previously reported associations between mLOY and factors such as age, smoking, all-cause mortality, and genetic loci.

The prevalence of mLOY in the UKB, as estimated by all three methods, significantly increases with age (Fig. 2A, Table 1). While the PAR-LOY method indicated the highest proportion of individuals with mLOY (around 27% at age 70), the strongest associations with age and smoking, as well as the best goodness of fit (Residual deviance), were observed with the MosCoverY estimate according to logistic regression analyses (Table 1).

**Table 1.**
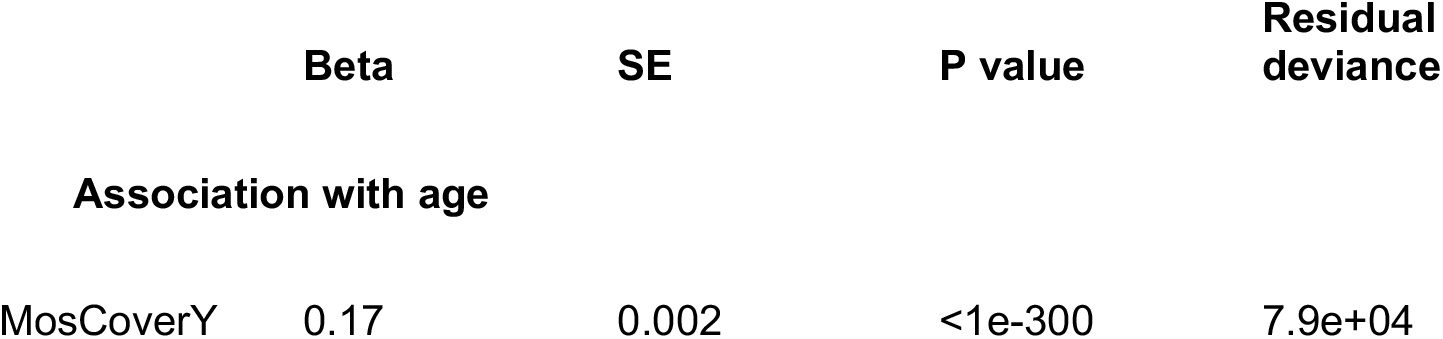
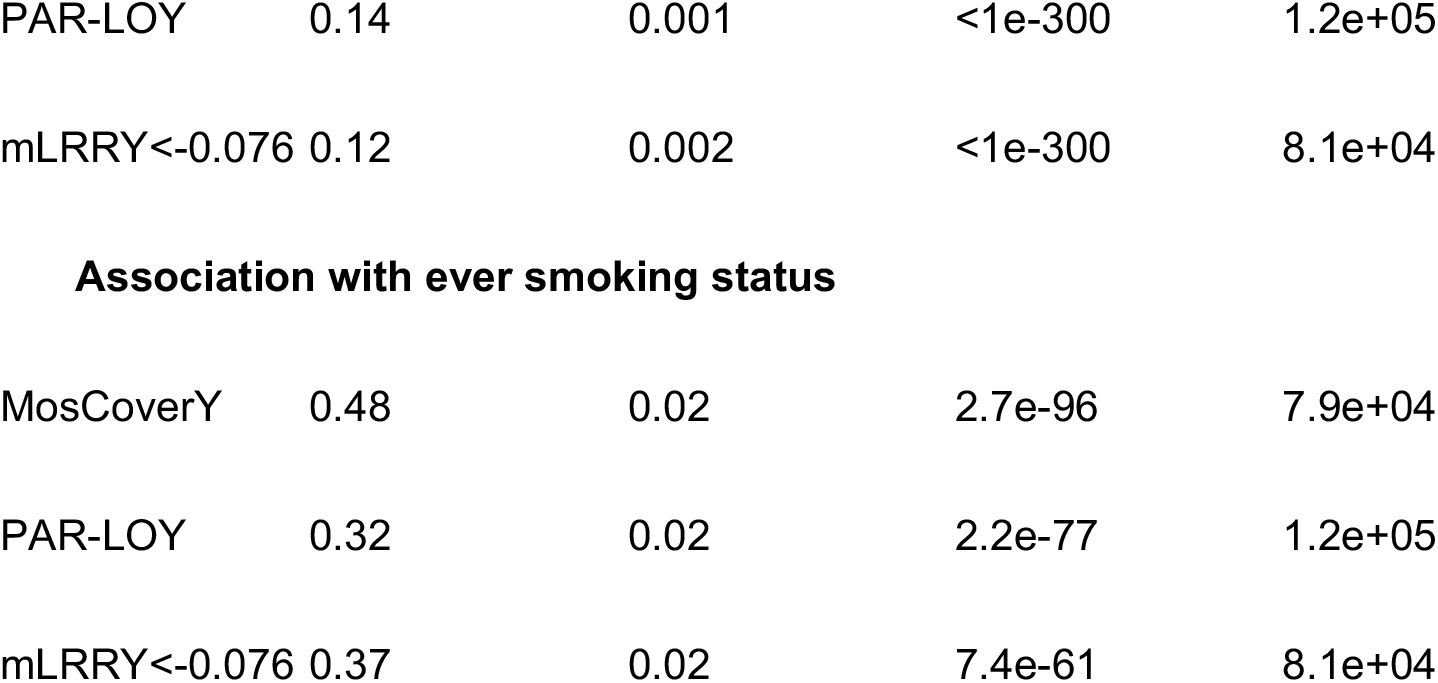
Association of mLOY binary trait estimated by three methods with age in smoking.

|  | **Beta** | **SE** | **P value** | **Residual deviance** |
| --- | --- | --- | --- | --- |
| **Association with age** |  |  |  |  |
| MosCoverY | 0.17 | 0.002 | <1e-300 | 7.9e+04 |

|  |  |  |  |  |
| --- | --- | --- | --- | --- |
| PAR-LOY | 0.14 | 0.001 | <1e-300 | 1.2e+05 |
| mLRRY<-0.076 | 0.12 | 0.002 | <1e-300 | 8.1e+04 |
| **Association with ever smoking status** |  |  |  |  |
| MosCoverY | 0.48 | 0.02 | 2.7e-96 | 7.9e+04 |
| PAR-LOY | 0.32 | 0.02 | 2.2e-77 | 1.2e+05 |
| mLRRY<-0.076 | 0.37 | 0.02 | 7.4e-61 | 8.1e+04 |

**Figure 2.**
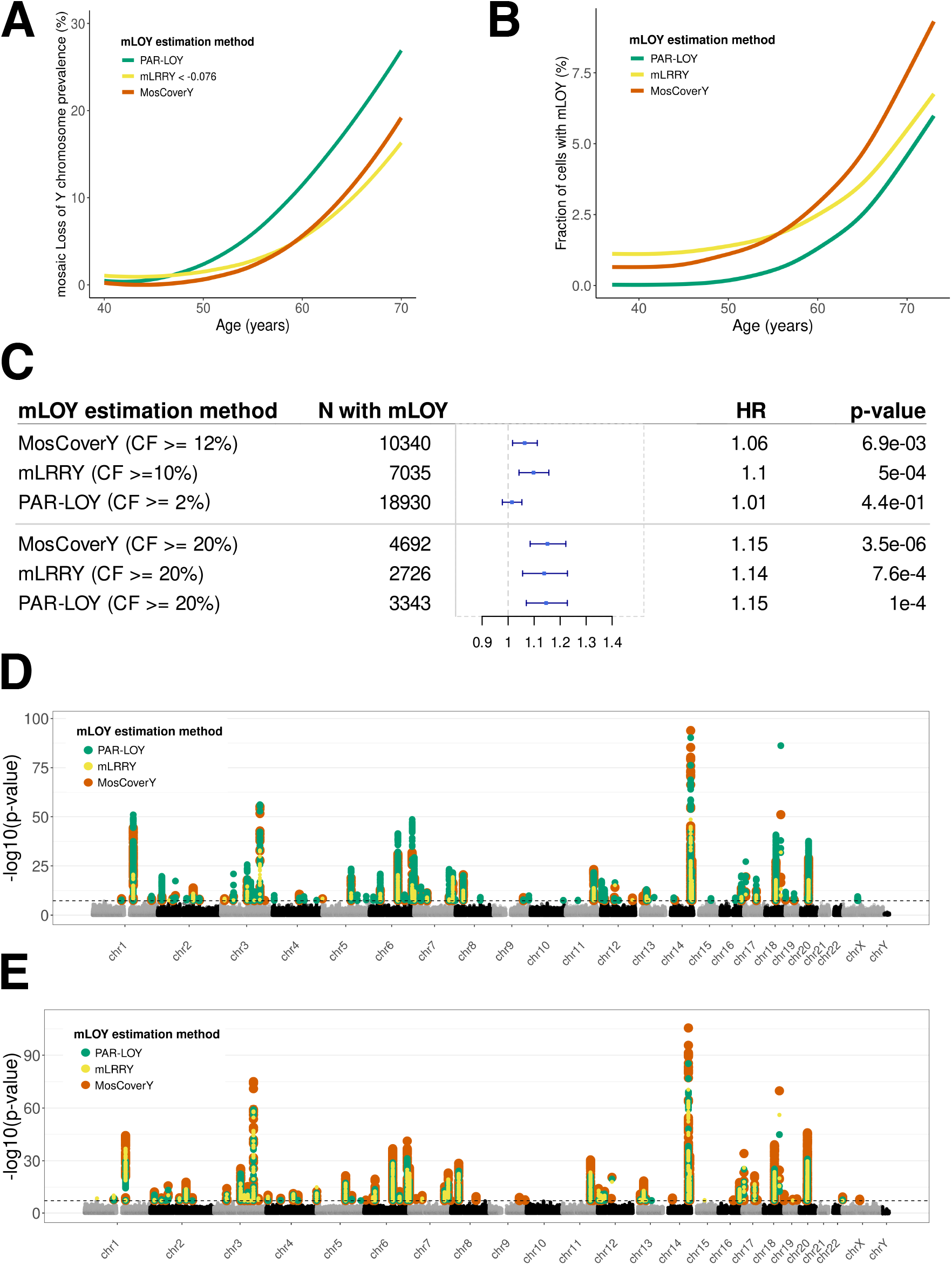
Biological validation of MosCoverY method. A - The prevalence of binary mLOY estimated by three methods increases with age. B - The fraction of cells containing mLOY estimated by three methods increases with age. C - Association of binary mLOY trait with all-cause mortality estimated by Cox proportional hazard model adjusted for age and smoking status. CF - fraction of cells with mLOY; HR - hazard ratio. D, E - Manhattan plot showing genetic loci associated with binary mLOY trait (D) and fraction of cells with mLOY (E). Each dot represents the genomic position, and the black horizontal dashed line represents a significant p-value threshold (5e-8).

The fraction of cells with mLOY was also positively associated with age and smoking (Fig. 2B, Table 2), with the MosCoverY estimates again showing the strongest association and highest R-squared value.

**Table 2.**
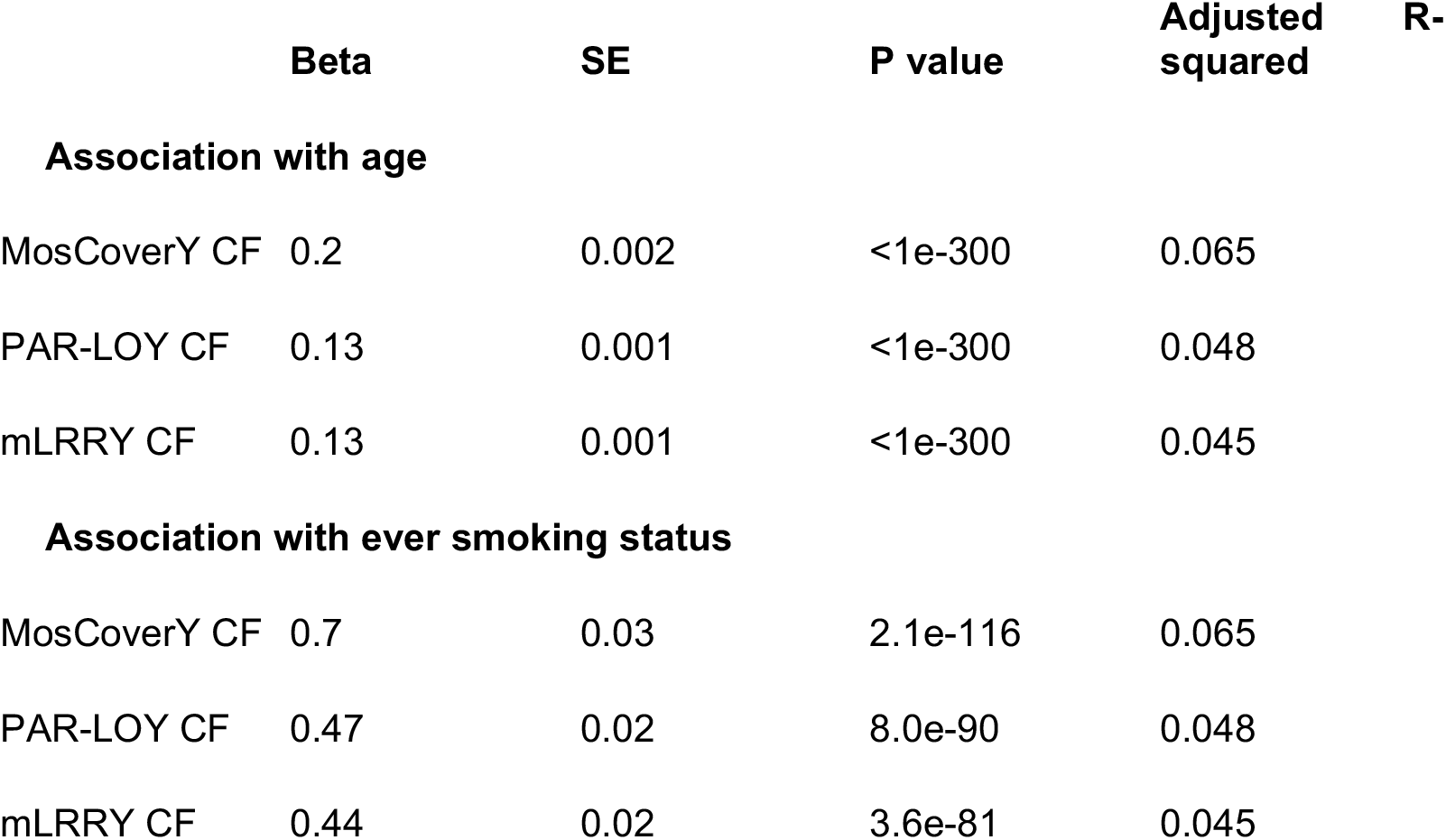
Association of fraction of cells (CF) with mLOY estimated by three methods with age in smoking.

We further assessed the previously reported association of mLOY with all-cause mortality (37). Using a Cox proportional hazards model, we evaluated time to death with mLOY binary traits as explanatory variables, adjusting for age, smoking status, and genetic principal components. Binary mLOY estimates from MosCoverY and mLRRY were significantly associated with all-cause mortality, with mLRRY showing a slightly stronger association. In contrast, the PAR-LOY estimates were not associated with all-cause mortality, likely because most mLOY carriers identified by PAR-LOY had mLOY in fewer than 10% of cells. Higher fractions of cells with mLOY are known to be associated with a greater risk of death (20). When defining the binary mLOY trait by a higher cell fraction (≥20%), we observed stronger associations with all-cause mortality across all three methods, with MosCoverY showing the strongest association (Fig. 2C).

In addition, we compared the association of mLOY with age and smoking across different groups of mLOY cases identified by one, two, or all three methods (as depicted in Fig.1C). The intersection of mLOY cases identified by all three methods (N=6,113) showed the strongest association with both age and smoking. Among the cases identified by two methods, the group identified by MosCoverY and mLRRY (N=1,362) showed the next strongest association. In the groups identified by only one of the methods, MosCoverY (N=1,252) demonstrated the strongest association with age and smoking, followed by PAR-LOY (N=11,440; Supplementary Table 3). When assessing association of mLOY with all-cause mortality in the same groups (but in the subset of European ancestry individuals), the only significant association was in the group of mLOY cases identified by all three methods (Figure S8).

### Genetic validation of MosCoverY

Previously, many genetic loci have been identified as associated with mLOY, estimated with mLRRY and PAR-LOY, through GWAS (10, 23). To better compare the strength of these known genetic associations for three mLOY estimates, we repeated GWAS on the set of 178,073 UKB participants of European ancestry for mLOY traits estimated by mLRRY and PAR-LOY, and compared the results to MosCoverY. We performed separate GWAS for the binary mLOY trait (Fig.2D) and for the fraction of cells with mLOY (Fig.2E).

We identified 36, 50 and 23 independent genetic signals for binary mLOY calls from MosCoverY, PAR-LOY and mLRRY, respectively. A total of 31 loci were shared between MosCoverY and PAR-LOY; 23 between MosCoverY and mLRRY and 22 between PAR-LOY and mLRRY. The strongest association identified by three methods (rs56349439 at chr14:101168739) had the smallest p-value for MosCoverY. All SNPs identified as independently associated are listed in Supplementary tables 4-9.

We identified 59, 35, and 38 loci associated with the fraction of cells with mLOY, as estimated by MosCoverY, PAR-LOY, and mLRRY, respectively: 34 were shared between MosCoverY and PAR-LOY, 36 betweenMosCoverY and mLRRY, and 32 between PAR-LOY and mLRRY. Most of the genetic signals associated with the fraction of cells with mLOY intersected with those associated with the binary mLOY trait, and most showed the strongest associations for MosCoverY estimates (Fig.2E).

## DISCUSSION

In this study, we developed MosCoverY, a novel approach to estimate mosaic loss of the Y chromosome (mLOY) from exome and genome sequencing data. This method provides an individual-level estimate of normalized median chrY coverage, which is used to derive a binary mLOY estimate and a quantitative measure of the fraction of cells with mLOY. We demonstrated the robustness of the MosCoverY method by comparing it to two widely used mLOY estimation methods based on genotyping array data - mLRRY and PAR-LOY. Our results demonstrate that MosCoverY provides an accurate and biologically meaningful estimate of mLOY, successfully overcoming the challenges of the Y chromosome structure by accounting for gene copy number, homology with X chromosome, and GC-bias.

MosCoverY showed a strong correlation with estimates from mLRRY and PAR-LOY. The highest number of people with mLOY was identified by PAR-LOY, which can be explained by the relatively low threshold for the fraction of cells with mLOY used by default by this method: 2% vs 10% and 12% for mLRRY and MosCoverY, respectively.

Defining the binary mLOY phenotype requires selecting a threshold, which we approached in a statistically defined manner. However, an alternative approach involves setting an arbitrary threshold based on a predefined fraction of cells with Y chromosome loss (Fig. S5). As the threshold increases, the proportion of individuals with mLOY identified by all three methods rises, relative to the total number of mLOY cases identified by any method. Setting a higher threshold for the fraction of cells with mLOY may be appropriate in certain contexts, as a higher fraction of cells affected by mLOY is likely to have a more pronounced impact on health-related outcomes (like all-cause mortality in Fig 2C).

The fraction of cells with mLOY also correlated well between three methods, with the highest correlation coefficient between MosCoverY and PAR-LOY and the lowest for PAR-LOY and mLRRY (Fig. 1D-E). MosCoverY also showed the highest correlation with Control-FRECC-estimated fraction of cells with mLOY from WGS data (Fig. 1G-I). Despite a good correlation, the MosCoverY estimated cell fraction was on average higher compared to the two other methods, which can mean either the overestimation of the fraction of cells with mLOY by MosCoverY or its underestimation by PAR-LOY and mLRRY. Both possibilities cannot be confidently ruled out. One of the steps in MosCoverY is the rescaling of the normalized chrY coverage to match the median of the distribution to the expected value of 0.5 which might be a source of overestimation. The argument against it is the fact that the correlation of the fraction of cells with mLOY with other methods when it’s estimated from rescaled coverage values improves compared to cell fraction estimated from the original normalized coverage of chrY.

We also validated MosCoverY by replicating known associations between mLOY and various clinical, epidemiological and genetic measurements. As expected, the prevalence of mLOY showed a massive positive correlation with age, and significant associations were observed with smoking status and all-cause mortality. These findings are consistent with previous studies, underscoring the relevance of our method. Notably, MosCoverY’s estimates showed the strongest associations with age and smoking, suggesting excellent sensitivity and robustness. Additionally, MosCoverY’s ability to identify germline genetic loci associated with mLOY, often showing stronger associations (especially for quantitative fraction of cells with mLOY rather than binary mLOY trait), further supports its accuracy and utility in genetic studies.

The limitations of the MosCoverY method include the requirement of high-quality exome sequencing data, and its accuracy may be affected by sample-specific variations in sequencing depth and quality. Additionally, while MosCoverY effectively normalizes coverage against matched autosomal exons, discrepancies in GC content or other biases could still impact the results. Importantly, the exon matching procedure may need to be repeated if another exome capture kit was used to generate the sequencing data. Future improvements could focus on refining the normalization process and validating the method across diverse populations and sequencing platforms. Finally, MosCoverY cannot be used to estimate mLOY in an individual sample, because the rescaling procedure requires the application of the method to multiple exomes to estimate the population median and the binary mLOY threshold.

MosCoverY can be directly applied to cohorts with exome and genome sequencing data, offering a valuable tool for large-scale epidemiological studies of mLOY, the most common type of somatic, age-related DNA change. mLOY is part of a bigger group of somatic DNA changes called mosaic chromosomal alterations (mCAs). Apart from mLOY, they include losses and gains of large parts or whole chromosomes, as well as copy-neutral loss of heterozygosity. All mCAs increase in prevalence with age and many have been shown to be associated with clonal hematopoiesis and various health-related conditions such as hematological cancers (38), solid tumors, diverse types of infections (4), and dementia (39).

Understanding the prevalence and impact of mLOY can provide insights into age-related diseases and male-specific health risks, and potentially help to explain the longevity gap between men and women as was proposed in several studies (14, 40, 41).

## Data Availability

UK Biobank data are available for research purposes upon project submission as described - https://www.ukbiobank.ac.uk/enable-your-research/apply-for-access.

https://github.com/ValeraKus/mLOY_Exomes

## ACKNOWLEDGEMENTS

This research was conducted using the UK Biobank Resource under Application #84415.

Funding for this works comes from EPFL, from the Swiss National Science Foundation (grant 197721), the French National Research Agency (grant GENVIR ANR-20-CE93-003) and the European Commission through the Horizon Europe project UNDINE (grant ID: 101057100).

