## Supplementary figures for "MosCoverY: a new method to estimate mosaic Loss of Y chromosome from NGS sequencing coverage data"

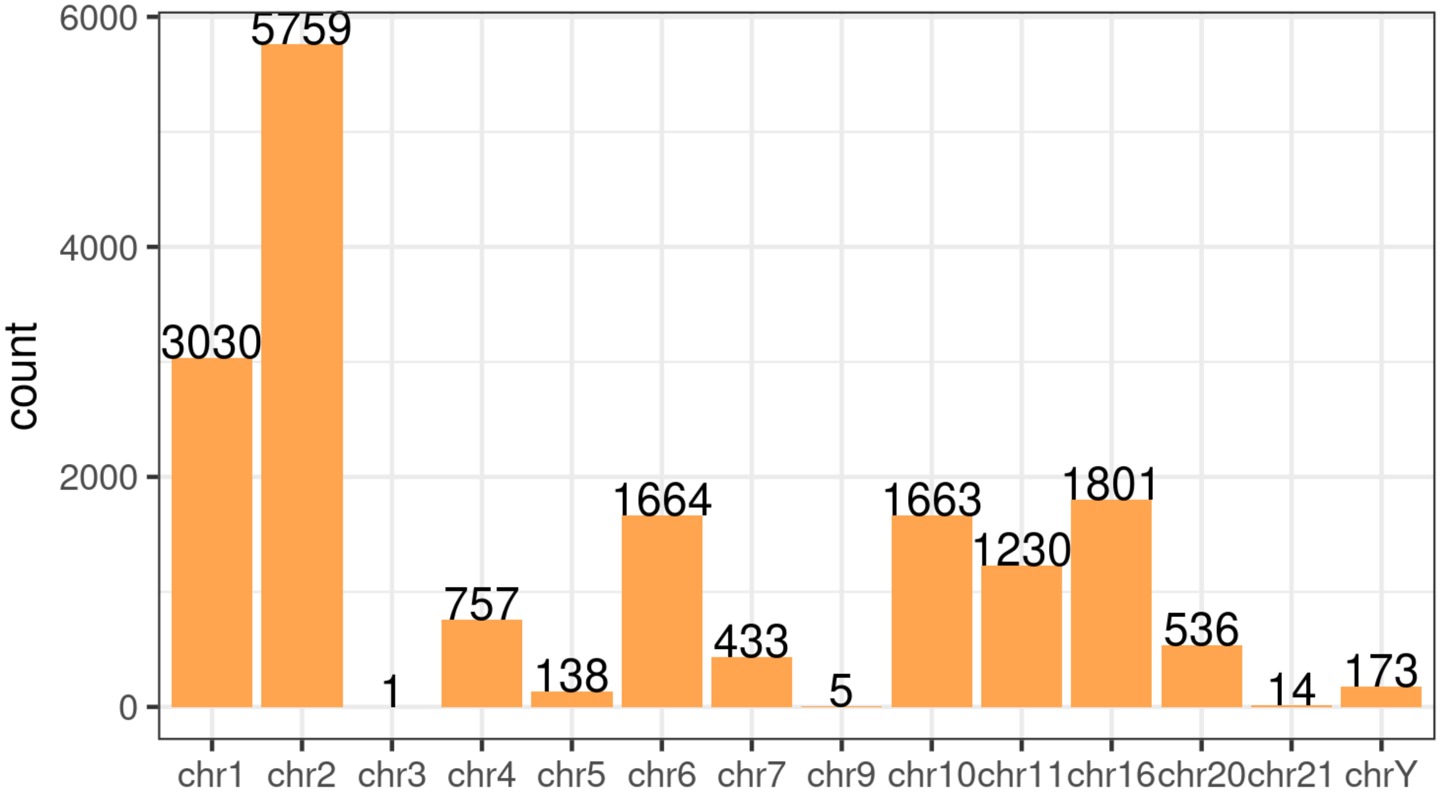

count
6000
4000
2000
0
5759
3030
1
757
138
1664
433
5
1663
1230
1801
536
14
173
chr1
chr2
chr3
chr4
chr5
chr6
chr7
chr9
chr10
chr11
chr16
chr20
chr21
chrY

Figure S1. Number of exons on each chromosome chosen for the MosCoverY: for each of the 173 exons on chrY we chose 100 exons for the coverage normalization on autosomes.

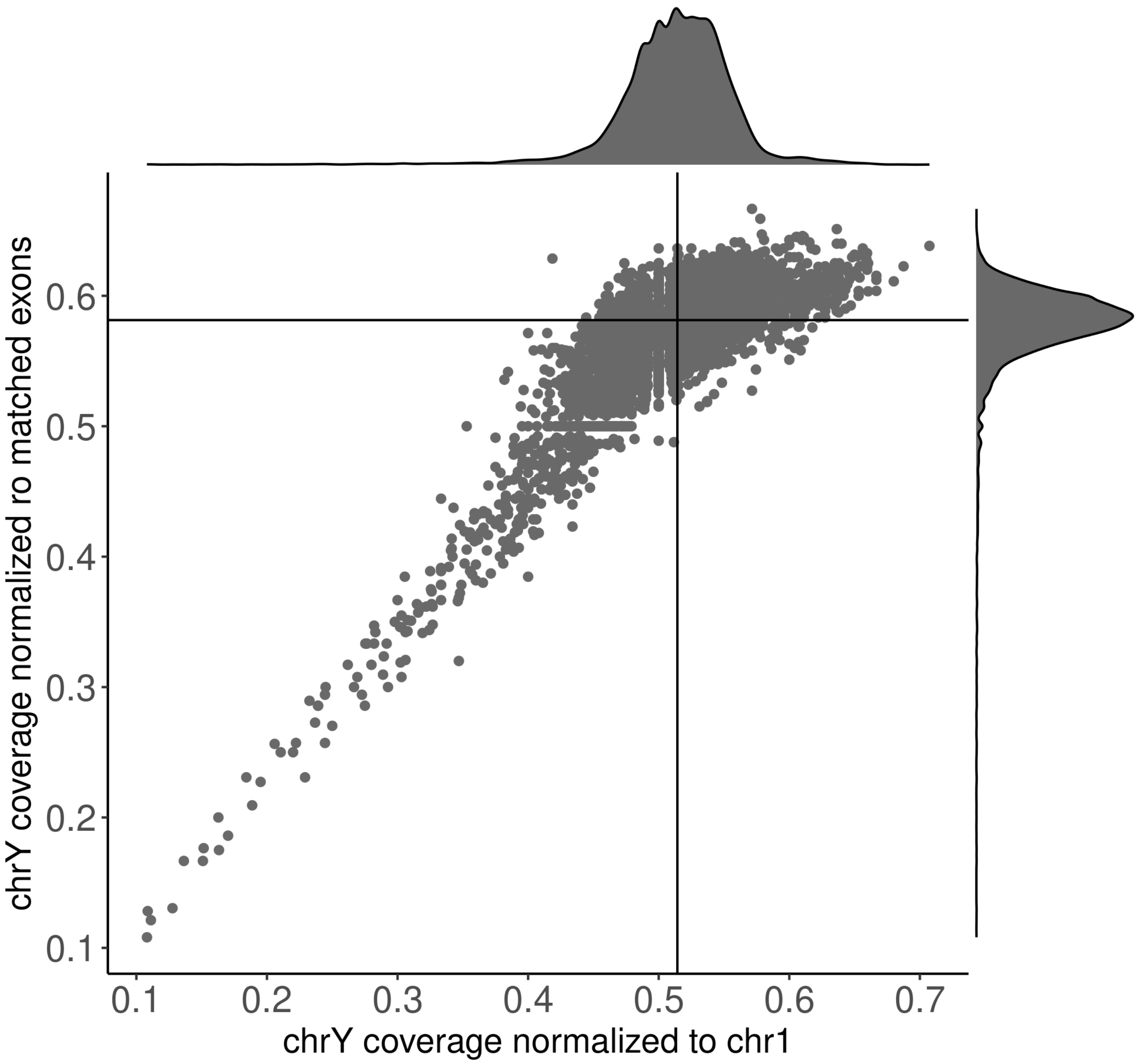

chrY coverage normalized ro matched exons
0.6
0.5
0.4
0.3
0.2
0.1
0.1
0.2
0.3
0.4
0.5
0.6
0.7
chrY coverage normalized to chr1

Figure S2. Comparison of individual-level estimate of normalized chrY coverage when normalizing on matched autosomal exons (Fig. S1) and on all exons on chr1 (N=10,000 randomly chosen individuals). The horizontal and vertical lines indicate the medians for the two estimates.

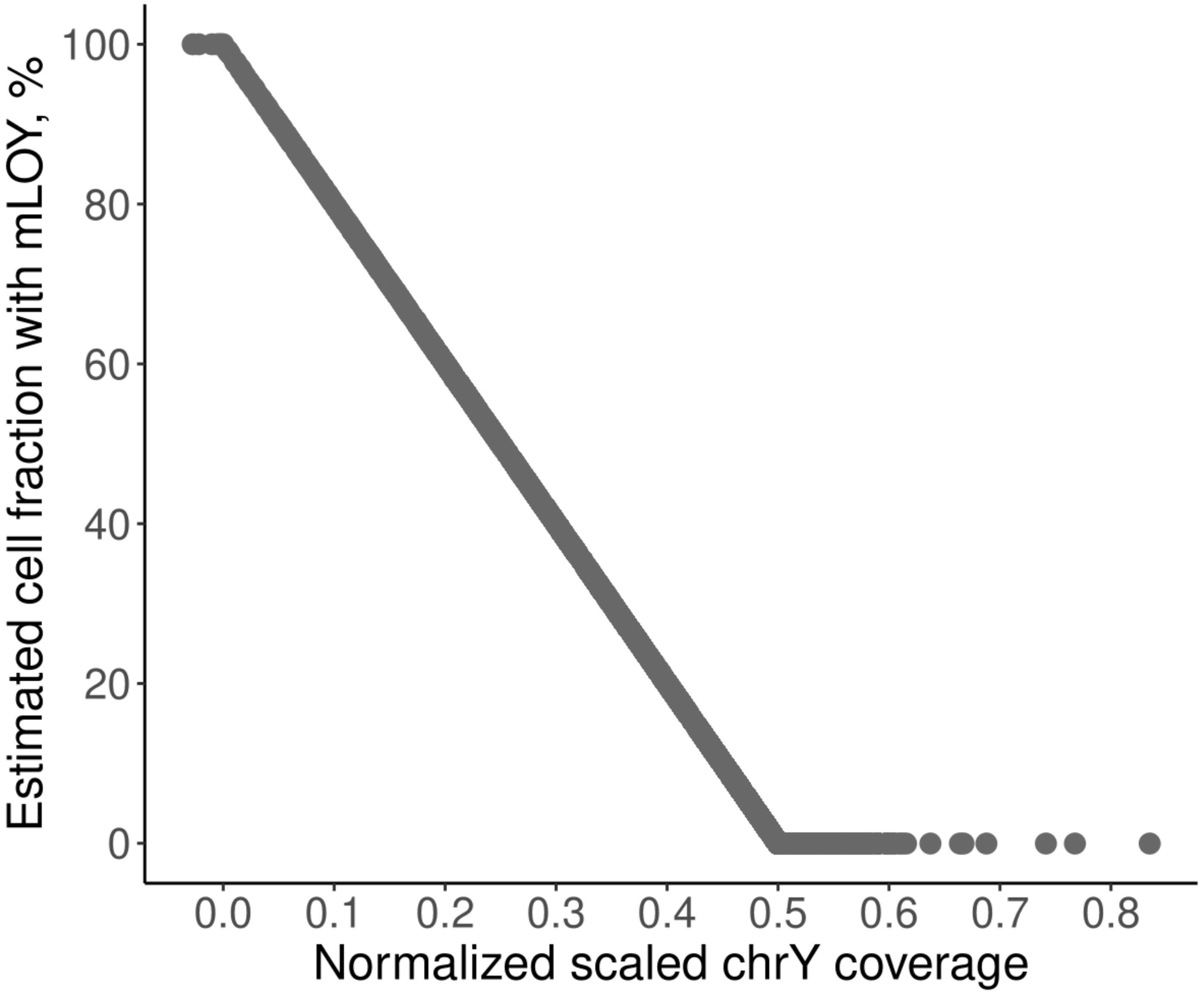
Estimated cell fraction with mLOY, %
100
80
60
40
20
0
0.0
0.1
0.2
0.3
0.4
0.5
0.6
0.7
0.8
Normalized scaled chrY coverage

Figure S3. Conversion of normalized scaled chrY coverage from MosCoverY to the fraction of cells with mLOY applying formula 1.

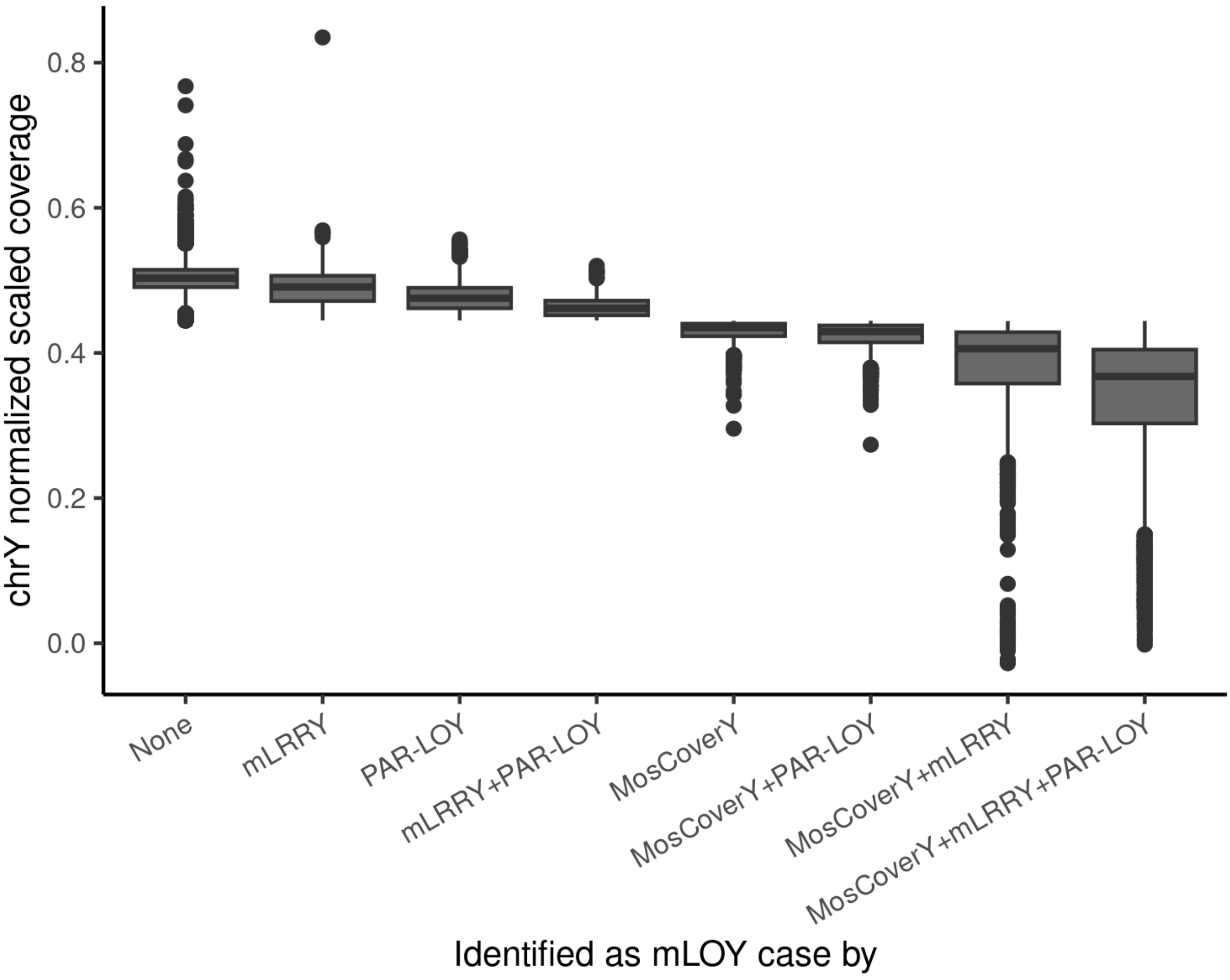

chrY normalized scaled coverage
0.8
0.6
0.4
0.2
0.0
None
mLRRY
PAR-LOY
mLRRY+PAR-LOY
MosCoverY
MosCoverY+PAR-LOY
MosCoverY+mLRRY
MosCoverY+mLRRY+PAR-LOY
Identified as mLOY case by

Figure S4. Normalized scaled coverage of chrY by the method of binary mLOY phenotype detection. For the number of individuals in each group see Fig. 1C.

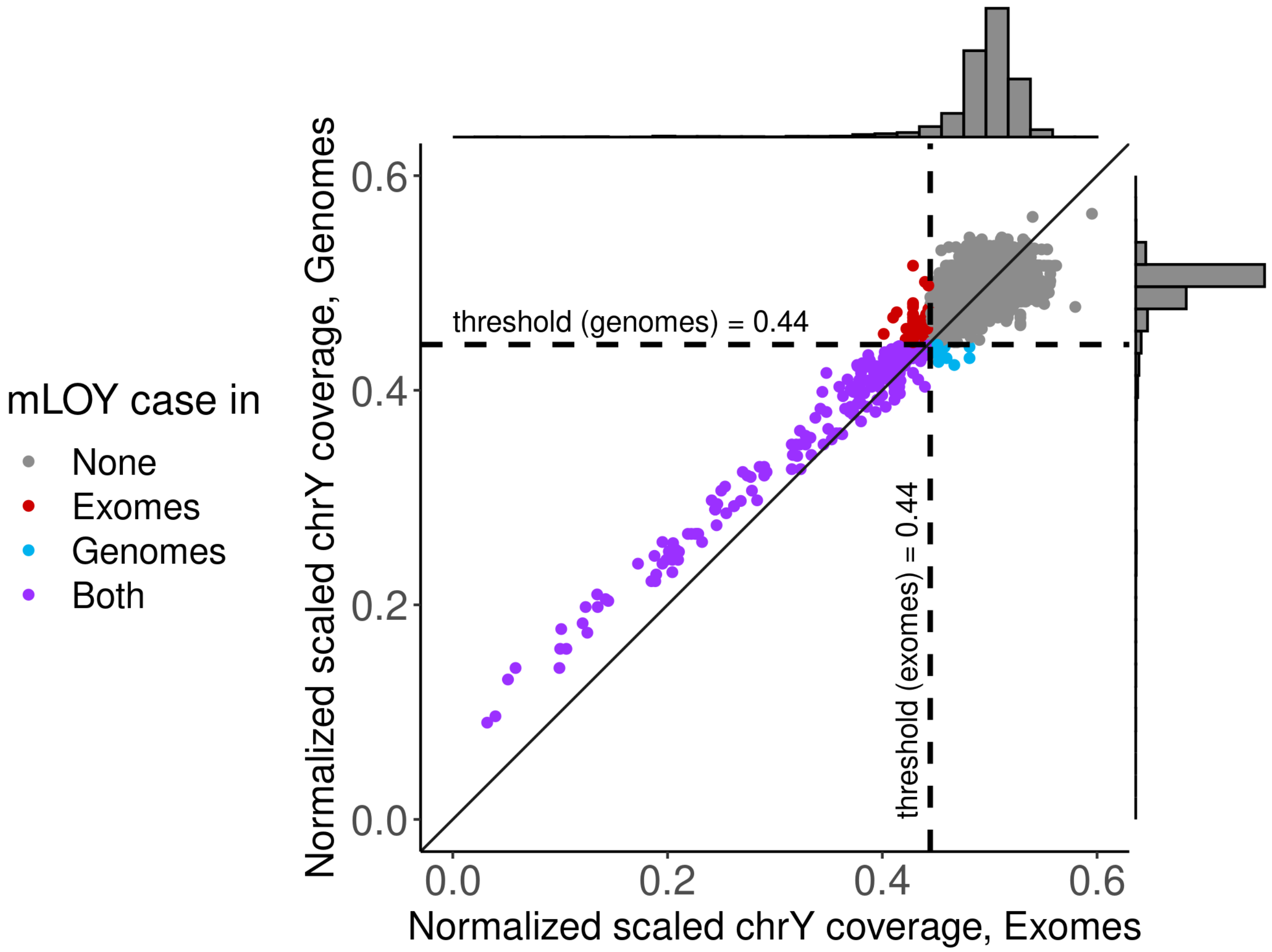
mLOY case in
None
Exomes
Genomes
Both
threshold (genomes) = 0.44
threshold (exomes) = 0.44
Normalized scaled chrY coverage, Genomes
Normalized scaled chrY coverage, Exomes
0.0
0.2
0.4
0.6

Figure S5. Comparison of applying MosCoverY to the exome and genome data, N=4,200. The thresholds are defined as Q1-1.5*IQR of normalized scaled chrY coverage for both genomes and exomes and showed as horizontal and vertical dashed lines.

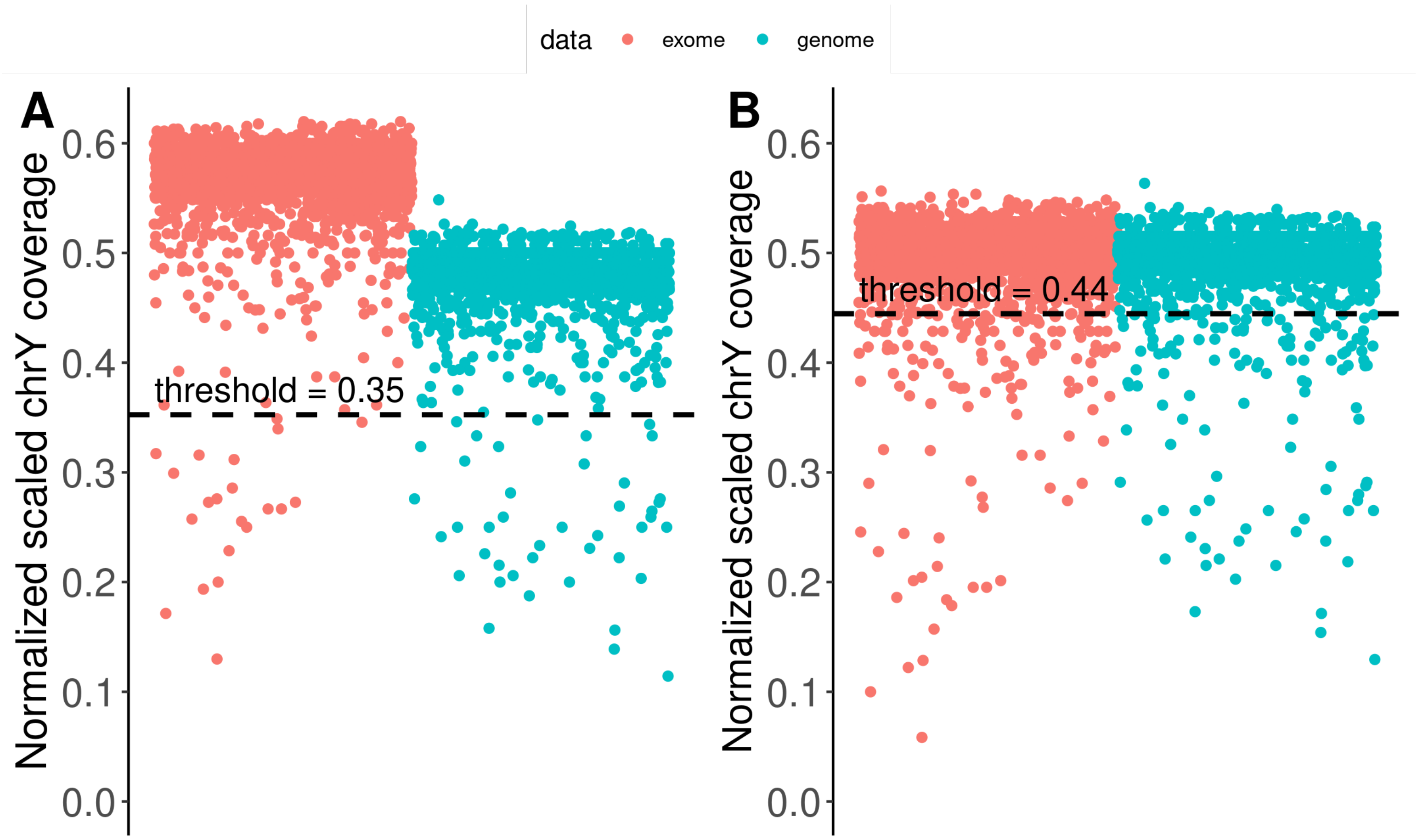

data
exome
genome
A
B
Normalized scaled chrY coverage
0.0
0.1
0.2
0.3
0.4
0.5
0.6
threshold = 0.35
threshold = 0.44

Figure S6. Estimating mLOY with MosCoverY from the mix of exome and genome data, N=4,200. A - exome and genome data are first combined and then rescaled to the population normalized chrY coverage of 0.5. B - exome and genome data are first rescaled to the population normalized chrY coverage of 0.5 and then combined. In both cases threshold is defined as Q1-1.5*IQR of normalized scaled chrY coverage.

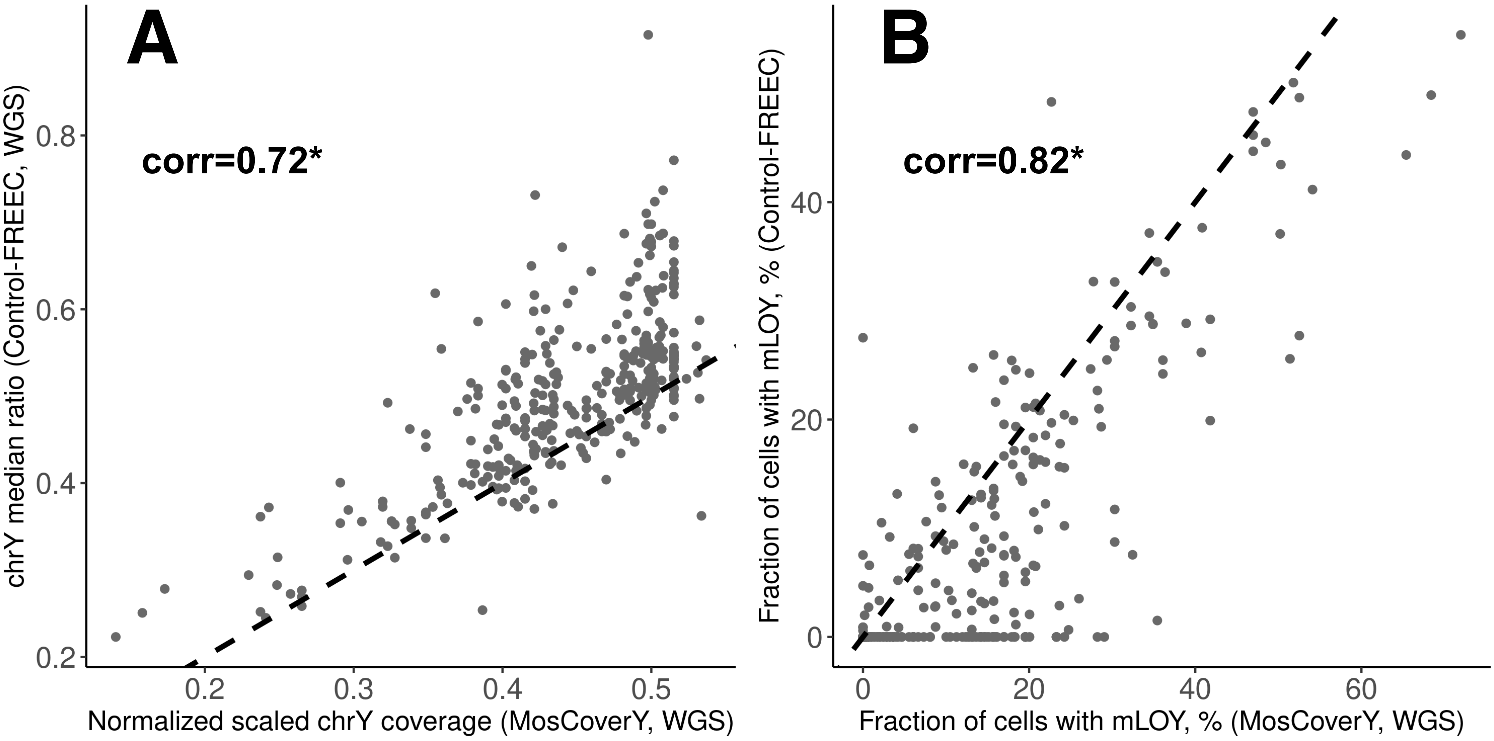

A
corr=0.72*
chrY median ratio (Control-FREEC, WGS)
0.2
0.4
0.6
0.8
0.2
0.3
0.4
0.5
Normalized scaled chrY coverage (MosCoverY, WGS)
B
corr=0.82*
Fraction of cells with mLOY, % (Control-FREEC)
0
20
40
0
20
40
60
Fraction of cells with mLOY, % (MosCoverY, WGS)

Figure S7. Comparison of MosCoverY results on WGS data and Control-Freec results for a subset of 360 randomly chosen individuals. A - Normalized scaled chrY coverage as estimated by MosCovery from WGS data vs chrY median ratio (normalized copy number) as estimated by Control-Freec from WGS data. B - Estimated fraction of cells with mLOY by MosCoverY applied to WGS data and Control-Freec (corr - Pearson's correlation coefficient, * - p-value ≤ 0.001).

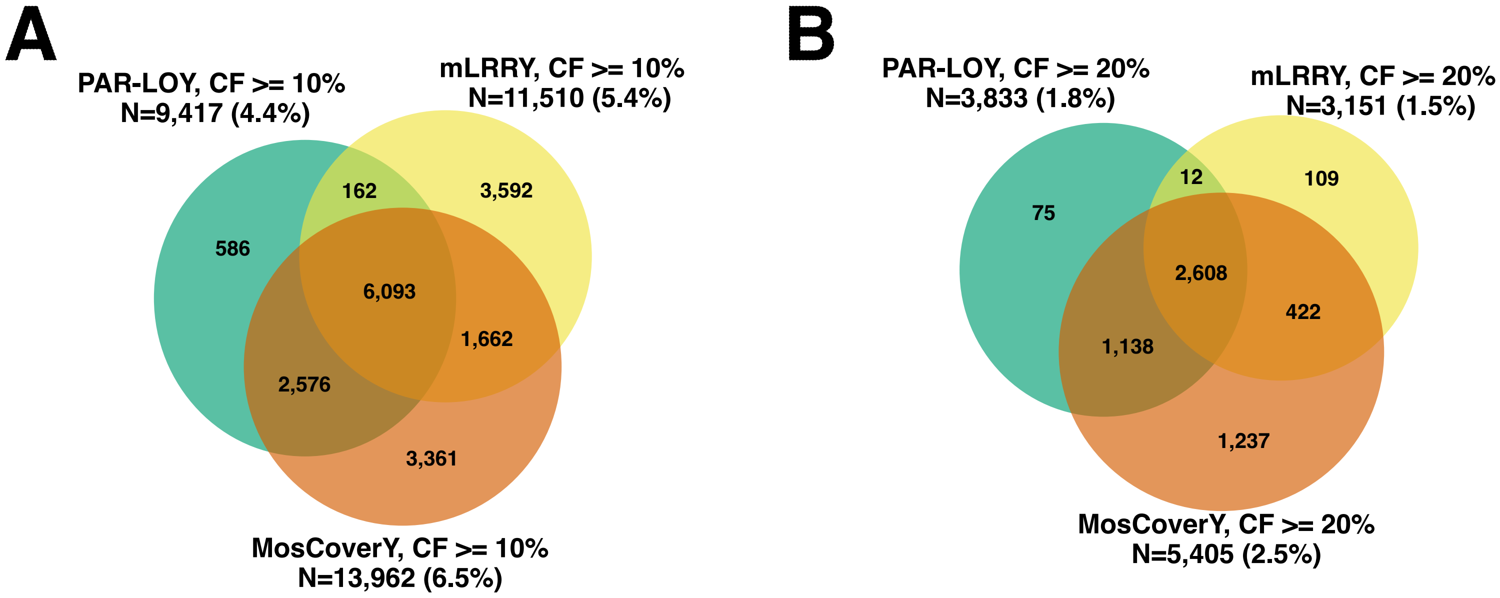
A
PAR-LOY, CF >= 10%
N=9,417 (4.4%)
mLRRY, CF >= 10%
N=11,510 (5.4%)
586
162
3,592
6,093
1,662
2,576
3,361
MosCoverY, CF >= 10%
N=13,962 (6.5%)
B
PAR-LOY, CF >= 20%
N=3,833 (1.8%)
mLRRY, CF >= 20%
N=3,151 (1.5%)
75
12
109
2,608
422
1,138
1,237
MosCoverY, CF >= 20%
N=5,405 (2.5%)

Figure S7. Comparison of different thresholds for defining binary mLOY estimates. A - Threshold at 10% of cells with LOY; B - Threshold at 20% of cells with LOY.

| mLOY estimation method | N with mLOY | HR | p-value |
|---|---|---|---|
| MosCoverY only | 1300 | 1.01 | 0.82 |
| PAR-LOY only | 10511 | 0.98 | 0.49 |
| mLRRY only | 1125 | 1.04 | 0.58 |
| MosCoverY and PAR-LOY | 3469 | 0.98 | 0.63 |
| MosCoverY and mLRRY | 960 | 1.11 | 0.11 |
| PAR-LOY and mLRRY | 339 | 0.84 | 0.21 |
| MosCoverY, PAR-LOY and mLRRY | 4611 | 1.12 | 0.00047 |

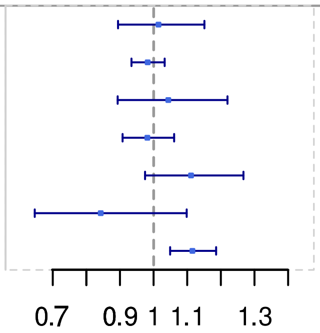

Figure S8. Association of binary mLOY traits defined by different combinations of methods with all-cause mortality estimated by Cox proportional hazard model adjusted for age and smoking status. Groups are defined by mLOY cases identified by one, two or all three methods. The analysis is performed on the individuals of European ancestry. HR - Hazard Ratio.
